# Determining Essential priorities for Future Investigation - a National consensus Exercise in Interventional Oncology (DEFINE-IO) – A priori protocol

**DOI:** 10.1101/2025.11.06.25339733

**Authors:** Vinson Wai-Shun Chan, Blair Graham, Helen Hoi-Lam Ng, Scott Griffiths, Deevia Kotecha, Hanif Ismail, Paul Jenkins, Jim Zhong, Raghuram Lakshminarayan, Tze Min Wah

## Abstract

**Introduction:** Interventional Oncology (IO), is a rapidly developing subspecialty within interventional radiology (IR), offering minimally invasive, precise and targeted cancer therapies. Despite its transformative potential, IO faces challenges in identifying critical research priorities amidst limited funding and resources. The DEFINE-IO study aims to identify the most pressing research priorities in IO to guide researchers, policy makers, and funding bodies in driving innovation and improving cancer outcomes.

**Methods and analysis:** This study comprises two phases: a modified e-Delphi process and a multi-criteria decision analysis (MCDA). The e-Delphi process will involve three rounds, with round 1 dedicated to generating research topics, followed by rounds 2 and 3, which will focus on refining and achieving consensus on the identified topics. The final consensus will yield a list of 25 research priorities, which will be evaluated during an in-person MCDA session. These priorities will be assessed against three predefined criteria: urgency, feasibility/equipoise, and affordability, to establish a strategic roadmap for research in IO.

**Ethics and dissemination:** Formal ethics approval was deemed not necessary. Findings of the study will be presented at major conferences and published in a peer-reviewed journal. The findings will also be shared with funding bodies and policymakers to help guide future research directions.

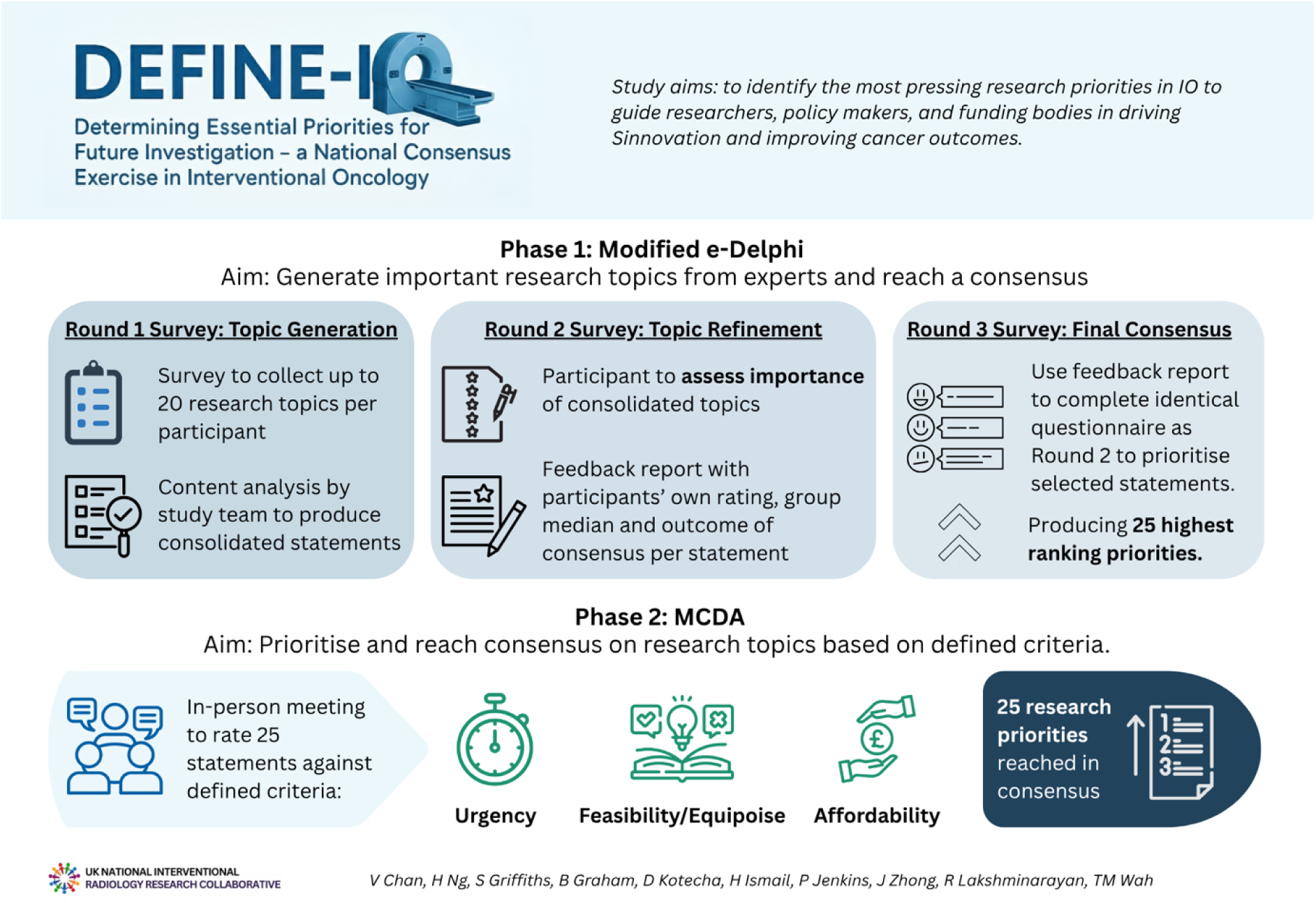

## 1. Introduction

Interventional Oncology (IO) is now established as the fourth-pillar of modern cancer care, offering a range of innovative, minimally-invasive therapies(1). Interventional oncologists utilise various techniques, broadly categorised into vascular and non-vascular approaches, to deliver precise, minimally invasive cancer therapies. Vascular IO leverages traditional vascular IR approaches via blood vessels in the body to perform procedures such as transarterial chemoembolisation (TACE)(2) and selective internal radiation therapy (SIRT)(3). These approaches are commonly used in the management of cancers such as hepatocellular carcinoma (HCC). Non-vascular IO performs a wide range of diagnostic, curative and palliative procedures, including biopsies(4), and image-guided ablation of thyroid(5), lung(6), liver(7), kidney(8) soft tissue(9) and bone malignancies(10).

Despite its transformative potential, research in the field often proceeds without a unified strategic direction(11). In a climate of limited research funding, there is a significant risk of fragmented efforts and the misallocation of precious resources towards questions that are not considered a priority by the clinical or patient community.

There is currently no formal consensus on the most critical research questions for IO in the UK. To ensure that future research in IO aligns with clinical needs and maximises its impact, it is essential to establish a clear framework of priorities. Therefore, the DEFINE-IO study is exceptionally timely as it directly addresses this critical gap. By applying a rigorous, evidence-based methodology utilising a modified Delphi technique(12) and a multi-criteria decision analysis (MCDA)(13–15), it will build the first national research strategy for the discipline.

Therefore, this study aims to identity the most pressing research priorities in IO to provide a critical roadmap to guide researchers, policymakers, and funding bodies in directing resources towards the most pressing clinical and scientific questions in IO.

## 2. Material and methods

### 2.1. Aims and objectives

The study’s main objective is to use a modified e-Delphi method to identify and prioritise research topics for interventional oncology. The secondary objective is to consider and reprioritise topics based on real-world deliverability.

### 2.2. Study design

This is a two-phase study integrating a modified e-Delphi method and a MCDA:

**Phase 1**: e-Delphi study to generate and reach consensus on important research topics.

**Phase 2**: MCDA to prioritise the agreed-upon research topics based on perceived urgency, feasibility/equipoise, and affordability implications.

#### 2.2.1. Rationale for modified Delphi method

The Delphi method is a well-established, systematic process for achieving expert consensus and has been used for research prioritisation in radiology(16), surgical oncology(17, 18), and emergency care(19). It involves multiple rounds of anonymous questionnaires with controlled feedback, promoting inclusivity and reducing bias by minimising the influence of dominant voices. Anonymity encourages honest responses, allowing participants to refine their views based on group feedback. Consensus is reached after 3 rounds.

In this DEFINE-IO study, a modified Delphi method will be adopted to identify research priorities, but real-world deliverability also depends on factors like urgency, feasibility/equipoise, and affordability. To address this, secondary analysis using MCDA will adjust the priorities to highlight those that are most impactful and achievable.

#### 2.2.2. Rationale for the MCDA

Following the modified Delphi assessment, MCDA will allow study participants to assess the deliverability of each research priority against defined criteria(14, 15). Proposed criteria include the urgency of addressing the priority, the feasibility of tackling the priority, and the associated cost and resource requirements. By applying weighted values to each criterion, it will be possible to adjust and rank the priorities for implementation in a practical, real-world context(20). While utilised in other contexts, utilisation of the MCDA is novel to research prioritisation and interventional radiology(13).

### 2.3. Scope of study

Research topics aligned with the European Curriculum and Syllabus for Interventional Oncology published by the Cardiovascular and Interventional Radiological Society of Europe (CIRSE)(21) will be included in the prioritisation exercise. Participants will be requested to suggest relevant research topics focusing on both technical and non-technical aspects of the delivery of interventional oncology.

### 2.4. Participants

#### 2.4.1. Population

Senior trainees, specialist doctors, and consultants with a substantive post in Interventional Radiology. Participation from the wider research team such as research nurses or radiographers are also encouraged.

To be included, participants must be at the level of specialty trainee year 5 (ST5) and above and be willing to participate in all study phases or be a research nurse or radiographer in interventional oncology.

Radiologists without a substantive IR commitment, and/or unable to commit to the study timeline will be excluded.

#### 2.4.2. Conflicts of Interest

To ensure transparency and avoid potential bias, all participants will be required to declare any potential conflicts of interest (COI) in advance. This includes any paid expenses or honoraria received from any relevant companies in the 12 months prior to the study. Recognising the limited pool of experts in this specialised field and the critical need to secure a sufficient and representative sample, participants declaring COIs will not be automatically excluded. Excluding these experts could introduce significant selection bias and compromise the validity and generalisability of the study’s findings.

Instead, this study will adopt a transparency-based management strategy. All declared COIs will be formally recorded. The aggregate number and nature of declared COIs will be reported in the final study. To assess the potential impact of these conflicts, a sensitivity analysis will be performed on the final results. Any substantive differences in the resulting consensus will be explicitly discussed as a limitation. The study oversight committee will review declared conflicts and monitor responses for any clear evidence of systematic bias.

#### 2.4.3. Incentives

Each participant will be provided with a certificate of participation upon completion of the final study questionnaire. PubMed indexed collaborator authorship will be offered to participants completing all three rounds of the modified Delphi study. No monetary incentive will be provided.

### 2.5. Sampling Strategy

A core participant group will be purposively recruited, aiming to ensure trainee and consultant representation from across 9 NHS regions and the three devolved nations. This includes (1) East of England, (2) London, (3) Midlands, (4) Northeast and Yorkshire, (5) Northwest, (6) Southeast, and (7) Southwest, (8) Wales, (9) Scotland, and (10) Northern Ireland. It is hoped that each region will contribute one IR trainee and one specialty doctor/consultant, yielding a minimum study sample of 20 participants.

Additional invites will be distributed via email and social media, using the study investigators’ existing networks. This includes the UK National Interventional Radiology Collaborative (UNITE), the British Society of Interventional Radiology (BSIR), and the Royal College of Radiologists (RCR). Invitations will promote involvement from women and groups traditionally under-represented within research.

Guided by available evidence to guide sample size in Delphi studies(22), recruitment will be stopped early once 60 participants are recruited.

### 2.6. Oversight Committee

A committee of study investigators and external representatives from UNITE, BSIR, and the RCR will oversee all stages of the study, including design, ethical considerations, questionnaire development, data analysis, and the dissemination of findings.

### 2.7. Study Process and procedures

#### 2.7.1. Study timeline

It is anticipated that data collection will run for five months from 1^st^ August 2025 to 7^th^ January 2026, as detailed in Table 1.

**Table 1:** Data collection timeline.

| Phase 1: e-Delphi Study |  |  |
| --- | --- | --- |
| Round 1 |  | Date |
| Participant Completion | 9 weeks | Commenced August 2025 |
| Data Analysis + Report | 1 week | Completed October 2025 |
| Round 2 |  |  |
| Participant Completion | 3 weeks | Commence October 2025 |
| Data Analysis + Report | 1 week | To be completed November 2025 |
| **Round 3 and Phase 2 MCDA** |  |  |
| Participant Completion (Hybrid Meeting with option to complete offline) | 4 weeks | To be completed January 2025 |
| Data Analysis + Report | 2 weeks | To be completed January 2025 |

#### 2.7.2. Survey Platform

A dedicated platform will be purpose built, with Microsoft forms and JISC Online Survey (https://onlinesurveys.jisc.ac.uk/) integrated for the Delphi study and the MCDA. This enables real time analysis of the Delphi results and implementation of the MCDA.

#### 2.7.3. Phase 1: e-Delphi

A three-round modified Delphi study is proposed consisting of topic generation, topic refinement and final consensus. For round 1, participants will each be invited to list up to 20 research topics or questions that they deem important for IO in the UK. Topics will then be consolidated by the study team using content analysis. The consolidated topics will be included within the round 2 questionnaire.

Round 2 will focus on topic refinement. Participants will be requested to assess the importance of each of the included topics against a forced-choice 9-point agreement scale, ranging from 1 (Extremely low priority) to 9 (Extremely high priority), outlined in Table 2(17). The median priority assigned to each topic by the participants will provide a measure of central tendency. A group median score of 1-3 indicates low priority, 4-6 indicates medium/ uncertain priority, 7-9 indicates high priority. Consensus will be determined using the IPRAS (Interpercentile Range Adjusted for Symmetry) as per the RAND/UCLA Appropriateness Method(23), with classical method as a sensitivity analysis(15). Findings from this round will be compiled into a feedback report, where each participant will see their own rating, the group median score, as well as whether consensus has been achieved for each statement. This report will be used to facilitate round 3.

**Table 2:** Forced-choice 9-point agreement scale; adapted from Schneider et al. (**17**)

| 1 | 2 | 3 | 4 | 5 | 6 | 7 | 8 | 9 |
| --- | --- | --- | --- | --- | --- | --- | --- | --- |
| Extremely Low Priority | Very Low Priority | Low Priority | Somewhat Low Priority | Medium priority | Somewhat High Priority | High Priority | Very High Priority | Extremely High Priority |

Round 3 will determine final consensus. Participants will be asked to repeat an identical questionnaire to that used during round 2, but with answers informed by the feedback report to inform their choices. Repeat analysis of this data, using identical methodology to that employed following round 2, will determine final priorities and consensus. In the event of an excessive number of statements, the selected statements that have exhibited uncertainty or a lack of consensus will be prioritised for the third round.

In rounds 2 and 3, the panellists will be asked to rate their expertise for each statement on a scale of no expertise, low expertise, moderate expertise, or high expertise. This will allow for sensitivity analysis to be performed based on the panellist’s expertise on each specific statement.

The Delphi study will provide a prioritised list of topics, ordered by the median rating and degree of consensus. The top 25 priorities that have achieved consensus, ranked by median and interpercentile range (IPR), will be subjected to analysis using multi-criteria decision analysis (MCDA).

#### 2.7.4. Phase 2: MCDA

An in-person meeting with a hybrid option will be planned to facilitate the MCDA process. This method provides a structured framework for making complex decisions by systematically evaluating various options against multiple, often competing, criteria. Reprioritising topics based on MCDA scores aims to augment Delphi findings by giving an indication of the real-world deliverability of each research topic. The MCDA will be conducted in a facilitated workshop setting to ensure all participants have a shared understanding of the process and to manage group dynamics. Participants from the e-Delphi phase will be invited to take part. The MCDA process will consist of three main stages: problem structuring, performance scoring and preference elicitation (weighting).

##### 2.7.4.1. Problem Structuring

The decision problem is to rank the top 25 research priorities (alternatives) based on their real-world deliverability.

- Alternatives: The 25 highest-ranking research topics identified from the e-Delphi process.
- Criteria: The alternatives will be evaluated against three predefined criteria. To ensure consistent interpretation, each criterion is defined as follows:

`o` Urgency: The degree to which the research topic addresses a pressing clinical or patient need that requires immediate attention. This considers the potential for the research to rapidly impact patient outcomes or resolve significant existing uncertainties in clinical practice.
`o` Feasibility/Equipoise: The likelihood that a research project on this topic can be successfully designed, executed, and completed within the typical constraints of the UK research environment. This includes considerations of patient recruitment, ethical approval complexity, the availability of required technical expertise and infrastructure, and whether there is equipoise for that investigation.
`o` Affordability: The estimated cost and resource requirements for conducting research on this topic. This is a ‘cost’ criterion, where a more affordable topic is considered more favourable. It encompasses not only direct financial costs but also demands on personnel, equipment, and facilities.

##### 2.7.4.2. Performance Scoring

Participants will assess the 25 research topics against each of the three criteria, populating a performance matrix.

- Scoring method: A constructed scale with direct rating will be used. For each criterion, participants will rate each of the 25 topics using a five-point Likert Scale with clearly defined performance levels to ensure consistency. The scale will be anchored as follows:

`o` Urgency: 1 (Very Low Urgency) to 5 (Very High Urgency)
`o` Feasibility/Equipoise: 1 (Very Low Feasibility/Equipoise) to 5 (Very High Feasibility/Equipoise)
`o` Affordability: 1 (Very High Cost/Low Affordability) to 5 (Very Low Cost/High Affordability)
- Normalisation: The raw scores from the Likert Scale will be converted to a common numerical value scale from 0 to 100, where 0 represents the worst possible performance and 100 represents the best. This normalisation allows for the meaningful aggregation of scores across different criteria.

##### 2.7.4.3. Criteria Weighting

In the first instance, each criterion will be assigned an equal weighting (i.e., 0.33). Presentation of data using spider charts will provide an intuitive means of presenting disaggregated data and may assist onward decision making and selection of topics (example—Figure 1)

**Figure 1.**
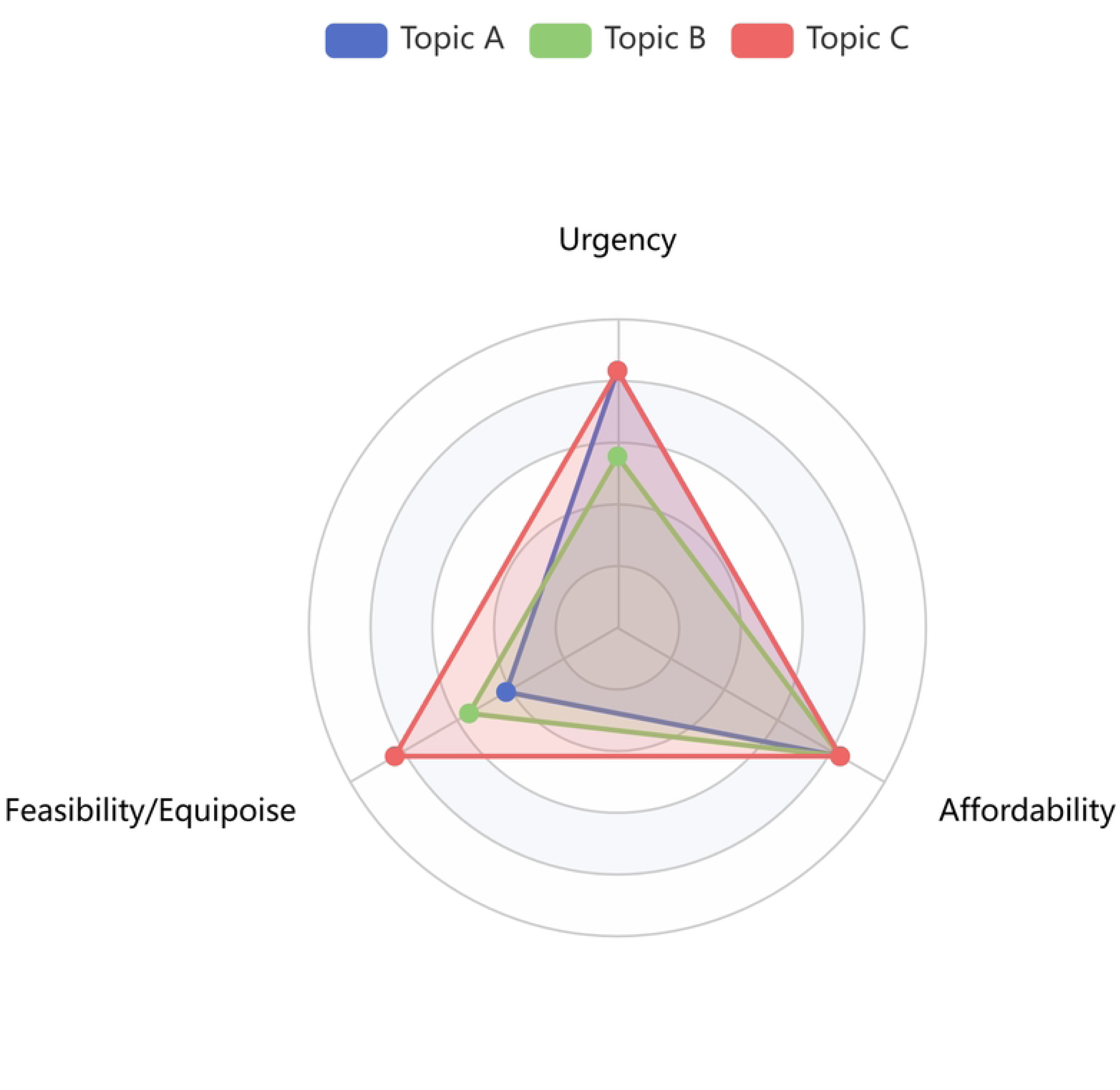
Example of how a spider chart may be used, following MCDA, to assess deliverability of topics based on urgency, feasibility/ equipoise and affordability. Whilst Topic A is most urgent, it is less feasible to investigate than Topic C. All three topics are equivalent in terms of affordability, and topic A and C are equivalent urgency.

To move beyond a simple assumption of equal importance, participants will elicit criteria weights that reflect their collective judgement on the relative importance of each criterion. The Swing weighing method will be used as a weighting method due to its theoretical robustness in capturing preference trade-offs(15). Prior to the session, a pilot process will be completed within the UNITE. The process is as follows(15):

1. Establishment of a Baseline Scenario: A baseline scenario describing a hypothetical research topic that performs at the lowest possible level on all three criteria will be presented to all participants. This establishes a common reference point for the weighting exercise.
2. Identification of the Primary Criterion: Participants will be asked the following question: “If you could change the performance of just one criterion from its worst level to its best level, which single change would provide the most significant improvement?”

- Each Participants will independently write down their selected criterion.
- A formal vote will be conducted to aggregate the individual selections. The criterion receiving the highest number of votes will be designated as the primary criterion.
3. Benchmark Scoring: The primary criterion identified in the preceding step will be assigned a benchmark score of 100 points. This score will serve as the anchor for the relative scoring of the remaining criteria.
4. Relative Scoring of Secondary Criteria: The remaining two criteria will be scored relative to the 100-point benchmark. For each criterion, participants will be asked: “Compared to the 100-point value assigned to the ‘swing’ of the primary criterion, how many points would you assign to the ‘swing’ of this criterion?”

- Each participant will privately record a score between 0 and 100 for each of the remaining criteria.
5. Data Aggregation: The scores for each criterion from all participants will be collected. The arithmetic mean of the scores for each criterion will then be calculated to produce a single, aggregate score.
6. Normalization of Weights: The aggregate scores will be normalized to create a set of weights that sum to 1.0.

Sensitivity analyses will be conducted to determine the effect of weighting priorities towards each of urgency, feasibility/ equipoise and affordability.

### 2.8. Data Analysis

#### 2.8.1. Content analysis for topic generation

The Delphi statements will be developed through a systematic thematic analysis of the initial, unstructured research suggestions provided by participants in round 1. To ensure a rigorous and clinically relevant structure, the analysis will be grounded in the framework of the CIRSE European Curriculum and Syllabus for Interventional Oncology(21). Initially, all suggestions will be reviewed to identify recurring topics and concepts. These will then be mapped onto the major sections of the CIRSE curriculum, a process that will allow for the establishment of several core research themes. Each individual suggestion will then be coded to an appropriate theme. Overlapping or related ideas will subsequently be synthesized and refined into distinct, unambiguous statements formulated to be clear, concise, and suitable for rating in a Delphi survey. This iterative process will ensure that the full spectrum of participant ideas, from broad concepts to highly specific procedural questions, is captured and logically organised.

#### 2.8.2. Descriptive Statistics

A comprehensive set of descriptive statistics will be calculated to summarise the data from each round of the Delphi study and the MCDA study. This analysis will go beyond basic descriptives to provide a detailed overview of the panel’s composition, engagement and emergent consensus.

The statistical summary will include but is not limited to participant characteristics including their professional role, years’ experience, geographic location, to describe the expert panel’s composition. Furthermore, response rates and engagement metrics will also be reported to ensure panel stability and engagement over time.

Following the thematic analysis of qualitative data from round 1, a descriptive summary of the emergent research topics will be presented. This will outline the key domains of the research identified by the panel.

For the Delphi study, basic descriptive statistics will include participant characteristics, survey characteristics at each round (including completion and time to completion for each survey), median priority assigned to each topic, and IPRAS for each item.

For the MCDA component, basic descriptive statistics will include survey characteristics, findings using equally weighted criteria, and sensitivity analyses. Presentation of findings using spider chart(s).

#### 2.8.3. Consensus Criteria (e-Delphi)

To assess group consensus and the relative priority of each research statement from Round 2 onwards, measures of central tendency (median priority score) and dispersion will be calculated. Consensus will be determined using the RAND/UCLA method, where a statement is deemed to have reached consensus if its interpercentile range (IPR) is smaller than its Interpercentile Range Adjusted for Symmetry (IPRAS). The IPRAS will be calculated for each item using the established formula: IPRAS = 2.35 + (1.5 * AI), where AI represents the asymmetry index(23).

#### 2.8.4. Sensitivity analyses

DEFINE-IO is a prioritisation exercise designed for the wider IO community, aiming to capture a broad and representative perspective across the field, therefore it is acknowledged that participants will naturally have differing levels of expertise in specific areas, but this diversity is an important part of reflecting the collective view of the IO community. However, it is acknowledged that some panellist may feel less confident in rating certain topics that they have no or less expertise in, hence a sensitivity analyse will be performed for the e-Delphi study with the participant’s self-perceived expertise. Furthermore, where necessary and feasible, sensitivity analysis will be performed based on the panellists’ year of experience, area of practice, and their baseline characteristics. Sensitivity analyses will also be performed in the MCDA utilising different weights.

### 2.9 Attrition Bias

To mitigate the risk of attrition bias, several strategies will be employed to maintain panel engagement. Regular and personalised communication will be maintained, and participants will receive summarised, anonymised feedback from the previous round to demonstrate the value of their contribution. Surveys will be designed to be as concise as possible to respect participants’ time. Furthermore, the characteristics of participants who complete all rounds versus those who withdraw will be compared to assess for any systematic differences.

## 3. Ethical Considerations

Formal ethical review and approval by an Institutional Review Board or Ethics Committee were deemed not necessary for the DEFINE-IO study. This decision is based on the nature of the research, which qualifies as a national consensus exercise focused on professional opinion elicitation rather than study on human subjects or clinical trial. Specifically, no human subjects research are involved. The participants are expert clinicians providing their professional judgment on future research priorities and deliverability, not contributing biological or personal data about themselves or patients. The study does not involve intervention, collection of clinical data, or testing of hypotheses related to health outcomes.

All panellists are provided with detailed information about the study’s purpose, procedures, risks, and benefits, with electronic written consent obtained prior to participation. Confidentiality is maintained by anonymising data and securely storing it in password-protected files, accessible only to the research team. Additionally, data protection measures ensure compliance with the UK General Data Protection Regulation (GDPR) and the Data Protection Act 2018.

## 4. Dissemination of Results

The study results will be compiled into a comprehensive report for participants and prepared for submission to peer-reviewed journals in interventional radiology. Findings will be presented at national conferences, such as the IOUK Annual meeting, BSIR Annual Scientific Meeting, and shared with funding bodies and policymakers to help guide future research directions.

## 5. Potential Limitations

Potential limitations include attrition between e-Delphi rounds and the MCDA phase, which may impact response rates. Additionally, there is a risk of subjective bias in scoring and weighting criteria. Overrepresentation of certain regions or institutions could skew the prioritisation of results. These limitations will be mitigated by ensuring a wide distribution of panellists from different regions and institutions, as well as minimising attrition by ensuring regular and effective communication with the study group.

## Data Availability

No datasets were generated or analysed during the current study. All relevant data from this study will be made available upon study completion.

## 6. Acknowledgements

None

## Notes

### Competing Interest Statement

I have read the journal's policy and the authors of this manuscript have the following competing interests: Dr Vinson Wai-Shun Chan has received research grants from Cancer Research UK and Leeds Hospital Charity, and has received support for travel from BVM Medical Ltd. Dr Helen Hoi-Lam Ng received support for travel from BVM Medical Ltd. Dr Jim Zhong has received a Cancer Research UK Clinical Trial Fellowship grant. Prof Tze Min Wah received research grant from HistoSonics, Johnson & Johnson, Boston Scientific and Angiodynamics as well as acting as consultant for Angiodynamics. The other authors have no competing interests.

### Funding Statement

Yes

